# Use of Stingless Bee Honey (Meliponinae Subfamily) in Wound Treatment: A Systematic Review and Meta-Analysis

**DOI:** 10.1101/2025.09.15.25335788

**Authors:** Deyziane Fernandes Da Silva, Felipe Silva Ferreira, Paulo C.C. do Nascimento, Alisson Luan S. de Assis, Kleyton Santos de Medeiros, Isabelle Katherinne Fernandes Costa, Rhayssa de Oliveira e Araújo

**Affiliations:** Graduate Program in Nursing, Federal University of Rio Grande do Norte, Natal, Rio Grande do Norte, Brazil; Onofre Lopes University Hospital, Brazilian Hospital Services Corporation (EBSERH), Natal, Rio Grande do Norte, Brazil; Department of Nursing, Federal University of Rio Grande do Norte, Natal, Rio Grande do Norte, Brazil; Teaching and Research Institute of the Northern Rio Grande do Norte League to Fight Cancer, Natal, Rio Grande do Norte, Brazil

## Abstract

**Introduction:** Stingless bee honey has been traditionally used in medicinal practices; however, its therapeutic properties have not yet been comprehensively systematized in the scientific literature. Honey produced by meliponine bees exhibits a unique biochemical composition that promotes wound healing, antimicrobial activity, and anti-inflammatory effects, particularly in chronic and hard-to-heal wounds such as those associated with diabetes mellitus—a significant public health challenge. This underscores the need for therapeutic alternatives that are effective, safe, and accessible.

**Objective:** To analyze the effectiveness of stingless bee honey in wound healing.

**Methods:** This is a systematic review with meta-analysis, to be conducted in accordance with PRISMA guidelines and registered in PROSPERO (CRD420251001075). Eligible studies will include clinical trials and in vivo or in vitro models that assess the use of pure stingless bee honey or its derivatives in wound healing. The intervention will be compared with standard treatments, placebo, or no treatment. Primary outcomes include wound size reduction, granulation tissue formation, and antimicrobial and anti-inflammatory activity. The search will be conducted in international databases with no restrictions on language or publication date. Two independent reviewers will perform study selection, data extraction, and risk of bias assessment.

**Conclusions:** Consolidating the existing evidence may contribute to a better understanding of the efficacy and safety of stingless bee honey in therapeutic wound care. The findings may support the development of new honey-based products and promote accessible treatment strategies, particularly for chronic wounds in resource-limited settings.

## 1. Introduction

The use of apicultural products for therapeutic purposes has been historically documented across various cultures. Honey, in general, is widely recognized for its wound-healing, antimicrobial, and anti-inflammatory properties ^[1,2]^. However, most existing studies have focused on the honey produced by *Apis mellifera*.

Stingless bee honey (Meliponini), commonly found in tropical regions, presents a distinct biochemical composition, characterized by unique attributes such as flavor, color, and viscosity, and holds therapeutic potential that remains underexplored ^[3,4]^. Brazil stands out for its vast biodiversity of stingless bee species, with over 300 identified types ^[5]^.

Although the majority of research on the therapeutic properties of bee products has centered on *Apis* honey, there is growing interest in the bioactive potential of stingless bee (Meliponini) honey ^[6,7]^. The chemical composition of these honeys is complex and varies depending on the bee species, local flora, and geographic region ^[7]^. Phenolic compounds, flavonoids, and organic acids are among the key components contributing to the bioactivity of these honeys ^[8]^.

Stingless bee honey exerts its effects through multiple mechanisms, influenced by its unique physicochemical properties such as high osmolarity, low pH, and hydrogen peroxide production. These factors contribute to its significant antimicrobial activity against both Gram-negative and Gram-positive bacteria ^[7,9,10]^.

Studies have shown that the use of stingless bee honey accelerates the wound healing process by maintaining a moist wound environment, which is crucial for cell migration and proliferation. It also stimulates angiogenesis and granulation tissue formation, thereby promoting re-epithelialization ^[1,11]^. Additionally, *Melipona subnitida* honey has been reported to increase collagen density, leukocyte infiltration, and fibroblast proliferation in infected wounds in rats ^[7,12]^.

Stingless bee honey also exhibits anti-inflammatory and antioxidant properties. The geopropolis of *Melipona fasciculata*, for instance, has been shown to reduce inflammatory cell infiltration in wounds of diabetic mice ^[13]^. Certain compounds present in the honey may protect cells from oxidative damage, which contributes to the healing process ^[14]^. Furthermore, honey also has debriding action, assisting in the removal of necrotic tissue and facilitating wound cleansing ^[15,16]^.

Despite growing public recognition and increasing scientific interest in Meliponini honey, significant knowledge gaps remain. Issues such as safety, standardization, appropriate methods of application, and robust evidence of efficacy in wound care still require more comprehensive and in-depth investigation ^[17]^.

Therefore, the central research question is: *What is the effectiveness of stingless bee honey in wound healing?* This review aims to analyze the efficacy of stingless bee honey in the healing of wounds.

## 2. Methods and Analyses

### 2.1 Registration

This review is registered in the PROSPERO database under the protocol number: CRD420251001075 ^[18]^. It will be conducted in accordance with the Joanna Briggs Institute (JBI) Manual for Evidence Synthesis and will follow the Preferred Reporting Items for Systematic Review and Meta-Analysis Protocols (PRISMA-P) guidelines ^[19]^.

### 2.2 Inclusion and Exclusion Criteria

Study eligibility for inclusion in this review will be established based on the PICOT framework (Population, Intervention, Comparators, Outcomes, and Time/Type of study), although the time component will not serve as a restrictive criterion.

- Population: Studies will be included if they involve animal models with experimentally induced wounds (e.g., rats and rabbits), *in vitro* models using cell cultures relevant to wound healing, or human subjects with wounds of various etiologies. In cases where the sample includes studies involving humans with different wound etiologies, results will be analyzed both collectively and separately, with a dedicated analysis for each etiology.
- Intervention: Studies that used pure stingless bee honey as the primary wound dressing will be selected. Included articles must report the bee subfamily or species, the dose, concentration, and method of honey application. Additionally, information on the floral source and/or geographic region where the honey was collected will be considered complementary but relevant. If studies using honey derivatives such as propolis or beeswax are identified, they will be analyzed separately.
- Comparators: Comparators will include standard treatments, placebo, or no treatment. Separate analyses will be conducted for studies comparing stingless bee honey with standard treatments and for those comparing it with placebo or no treatment.
- Outcomes:The primary outcomes to be assessed include wound healing time, changes in wound area (measured in square centimeters), and cell proliferation (evaluated through direct counting of fibroblasts and keratinocytes, or by assays such as MTT/XTT/WST, BrdU/EdU, Ki-67, or others). Granulation tissue formation will also be evaluated, based on visual assessment of the wound bed using specific scoring systems, quantification of newly formed tissue in mm^2^, µm^2^, or as a percentage of wound closure, as well as vascular density and cellularity determined through histological analysis.

Secondary outcomes will include the antimicrobial properties of honey, assessed using quantitative methods such as inhibition zone diameter (mm), minimum inhibitory concentration (MIC), minimum bactericidal concentration (MBC), colony-forming unit (CFU) count (log_10_), or bacterial kill time.

Anti-inflammatory activity will also be considered, evaluated through histopathological analysis, including microscopic observation of the intensity and type of inflammatory infiltrate (presence of leukocytes, polymorphonuclear and mononuclear cells), quantification of pro-inflammatory cytokines and other inflammation-related molecules, as well as assessment of classical signs of inflammation (pain, heat, redness, and edema), measured using quantitative scales.

- Timeframe / Type of Study: Eligible studies will include controlled laboratory experiments, both *in vitro* (cell cultures) and *in vivo* (animal models), as well as controlled experimental studies conducted in humans. There will be no restrictions regarding the duration of follow-up or intervention. Studies may be published in any language and from any time period.

The decision to include *in vitro, in vivo*, and human studies was based on the limited number of studies specifically addressing the use of stingless bee honey for wound healing in humans. Analyses of *in vitro, in vivo*, and human studies will be conducted separately, depending on the availability and compatibility of outcome variables.

Studies involving animals with pre-existing conditions or diseases that could directly interfere with the wound healing process (e.g., severe metabolic disorders or uncontrolled infections) will be excluded, unless such conditions are explicitly part of the study objective. Studies that do not specify the origin or composition of the honey used will also be excluded—for example, lack of identification of the stingless bee species, or absence of information regarding honey purity, processing, or chemical composition.

Studies in which the primary intervention does not involve honey from bees of the *Meliponinae* subfamily will be excluded, as will those using pasteurized honey (since pasteurization can degrade heat-sensitive bioactive compounds). Studies that apply honey in combination with other products that could alter or obscure the interpretation of results, as well as studies lacking quantifiable outcome data, will also be excluded.

Case reports or case series, theoretical studies or literature reviews, studies with incomplete data or no clear methodology, studies with a high risk of bias, poor methodological quality, or non-reproducible results will not be included in the final sample. Methodological quality will be assessed using the GRADE approach and the SYRCLE Risk of Bias Tool (for animal studies).

### 2.3 Search Strategy

The search will be conducted in the following databases: Medline, Elsevier’s Scopus (SCOPUS), Embase, Cochrane Central Controlled Trials Registry (CENTRAL), Web of Science, Latin American and Caribbean Health Sciences Literature (LILACS), SciELO, Cumulative Index to Nursing and Allied Health Literature (CINAHL), Science Citation Index Expanded (SCI-Expanded), and Science Direct.

The search strategy will be adapted for each database, according to its specific indexing and search functionalities (Table 1).

**Table 1.**
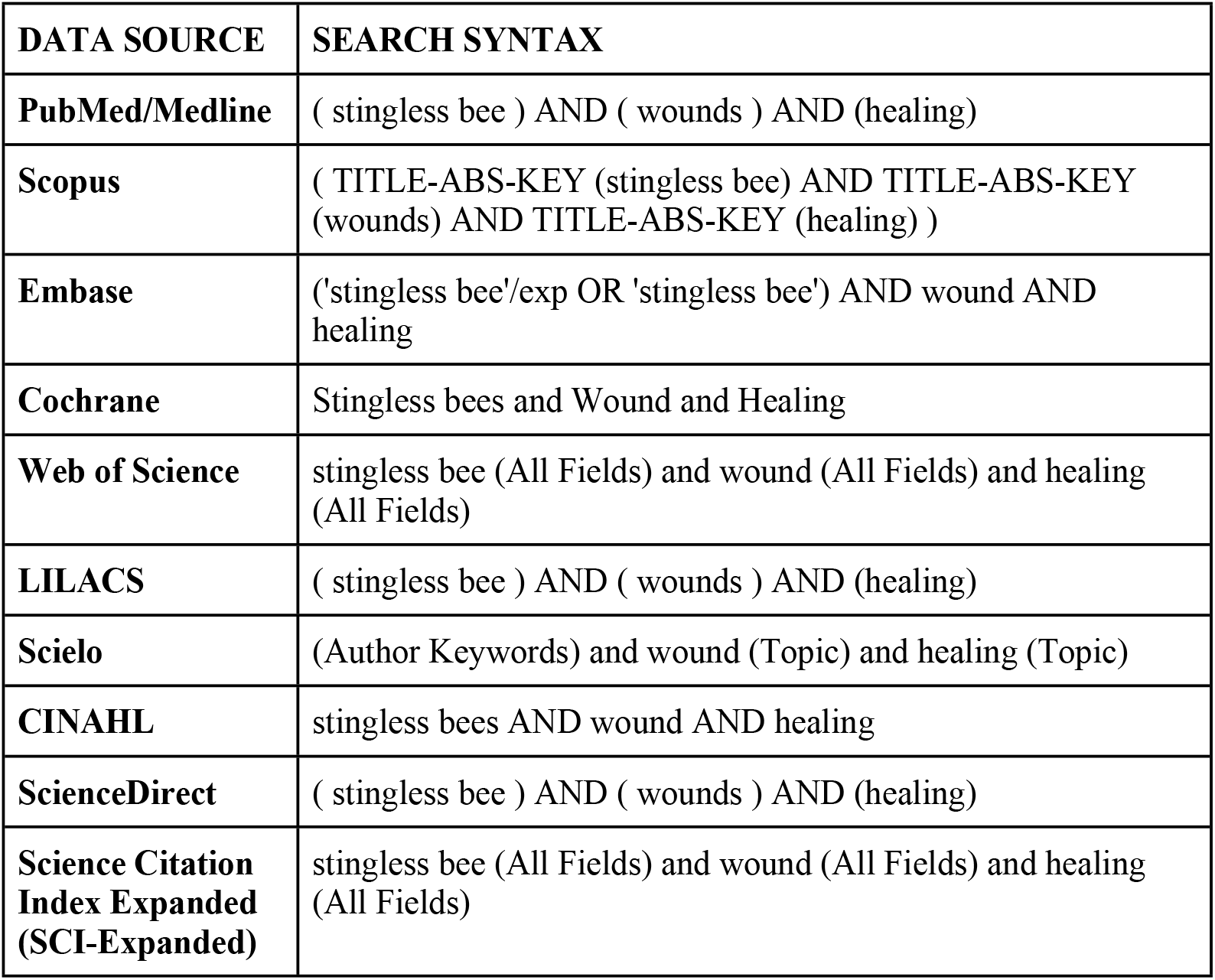
Search Strings by Database.

### 2.4 Study Selection

Two independent reviewers will screen the titles and abstracts, followed by full-text reading. Any disagreements will be resolved by a third reviewer. Data will be collected using a form specifically designed for this review. Prior to data collection, the reviewers will undergo standardized training to ensure a consistent understanding of the search criteria and proper use of the data extraction form.

Data collection will be carried out independently and in a blinded manner by two reviewers to minimize errors and bias. After data extraction, the results from both reviewers will be compared using the Rayyan software to identify discrepancies. In cases of disagreement, a third reviewer will be consulted to resolve the conflicts.

The reference lists of the included articles may also be used to expand the scope of the computerized literature search and identify additional eligible studies.

The detailed flow of the search and study selection process will be presented in a flow diagram, as shown in Fig. 1.

**Fig 1.**
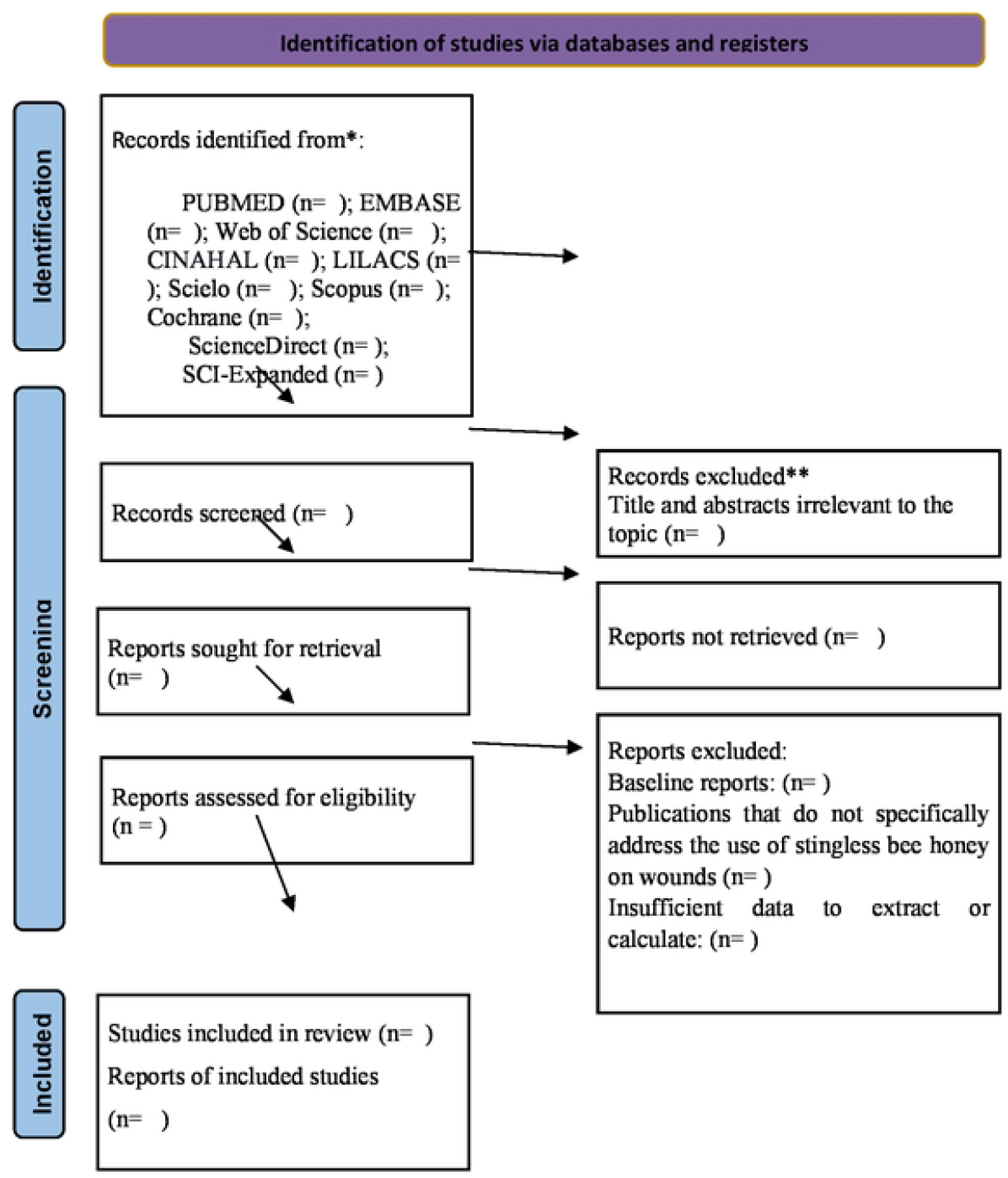
PRISMA flow diagram for systematic review and meta analysis.

### 2.5 Data Extraction

The following data will be extracted from the selected articles:

- Last name of the first author and year of publication;
- Country where the study was conducted;
- Study design;
- Wound healing time;
- Last name of the first author and year of publication;
- Country where the study was conducted;
- Study design;
- Wound healing time;
- Change in wound area (in square centimeters);
- Cell proliferation (assessed by direct counting of fibroblasts and keratinocytes, or by methods such as MTT/XTT/WST, BrdU/EdU, Ki-67, or others);
- Granulation tissue formation (measured through visual assessment of the wound bed using specific scoring systems, quantification of newly formed tissue in mm^2^, µm^2^, or as a percentage of wound closure, as well as vascular density and cellularity assessed by histological analysis);
- Antimicrobial activity (inhibition zone diameter in mm, minimum inhibitory concentration [MIC], minimum bactericidal concentration [MBC], colony-forming unit [CFU] count in log_10_, or bacterial kill time);
- Anti-inflammatory activity (measured through histopathological analysis, including microscopic observation of the intensity and type of inflammatory infiltrate—presence of leukocytes, polymorphonuclear and mononuclear cells—quantification of pro-inflammatory cytokines and other inflammation-related molecules, as well as classic signs of inflammation such as pain, heat, redness, and edema, using quantitative scales);
- Bee subfamily or species;
- Dose and concentration of stingless bee honey used;
- Frequency of stingless bee honey application;
- Method of stingless bee honey application;
- Floral source and region of honey collection (if available);
- Animal model used in the study (e.g., rabbit, mouse, or others) or human model;
- Potential adverse effects observed during the study.

After the data collection and verification phase is completed, the authors will conduct an in-depth analysis of all included studies. Forest plots will be generated for each study. Following data extraction, a meta-analysis will be performed for those studies that present the necessary characteristics for such analysis.

### 2.6 Risk of Bias Assessment

The risk of bias of the included studies will be independently assessed by two reviewers. In case of discrepancies, these will be resolved by consensus. If necessary, a third reviewer will be consulted to reach a final decision.

For preclinical animal studies, the SYRCLE Risk of Bias Tool (Systematic Review Centre for Laboratory Animal Experimentation) will be used. This tool is based on the original Cochrane Risk of Bias tool and adapted for animal experiments. It evaluates the following domains: random sequence generation, allocation concealment, blinding of investigators and outcome assessors, incomplete outcome data, selective outcome reporting, and other potential sources of bias.

For randomized clinical trials, the Cochrane Risk of Bias 2.0 (RoB 2) tool will be used. This tool evaluates five key domains: bias arising from the randomization process, deviations from intended interventions, missing outcome data, measurement of the outcomes, and selective reporting.

Each domain will be rated as having low risk of bias, some concerns, or high risk of bias, according to the criteria established by the respective tools. The results will be presented descriptively and, where applicable, visually through risk of bias graphs.

### 2.7 Assessment of Methodological Quality of the Studies

This systematic review will employ the GRADE Pro (Grading of Recommendations Assessment, Development and Evaluation) approach to assess the quality of the evidence supporting our findings. GRADE classifies the certainty of evidence into four levels: high, moderate, low, or very low, based on five key domains: risk of bias, inconsistency, indirectness, imprecision, and publication bias.

To ensure objectivity and minimize bias, two independent reviewers will conduct the GRADE assessment for each included study. Any disagreements or conflicts that arise between the two reviewers during the evaluation process will be resolved by a third reviewer.

### 2.8 Data Summary

The data will be described narratively and presented in tables summarizing the characteristics of the studies included in the sample.

A meta-analysis will be conducted for studies that present appropriate methodological characteristics and similar research questions, enabling data comparability and statistical validity. Data analysis will be performed using Review Manager (RevMan) software, version 5.4.

Selected studies must provide numerical data that can be extracted and used for statistical calculations. To minimize bias and ensure comparability of results, studies must use standardized methods for outcome measurement and diagnostic criteria for participant inclusion, as well as adopt similar follow-up periods. Studies with a high risk of bias may be excluded from the meta-analysis or analyzed separately.

### 2.9 Heterogeneity

The chi-square (χ^2^) test will be used to assess the consistency of the results across the included studies, with a significance level set at *p* < 0.1 [20]. Heterogeneity will also be evaluated based on the criteria described in the Cochrane Handbook, using the I^2^ statistic. This test quantifies the proportion of variability across studies that can be attributed to true heterogeneity rather than chance. The I^2^ statistic provides a numerical measure of inconsistency between studies, supporting the interpretation of the magnitude of heterogeneity.

A random-effects model will also be used, given its suitability for high heterogeneity. Unlike the fixed-effect model—which assumes that all studies share a single true effect—the random-effects model is more appropriate when substantial heterogeneity is present, as it accounts for variability between studies, allowing each study to have its own true effect size.

For this protocol, the interpretation of I^2^ will follow the following thresholds: 0% to 25% – low or no heterogeneity; 26% to 50% – moderate heterogeneity; 50% – substantial or high heterogeneity.

It is important to note that this assessment will only be conducted if a meta-analysis is deemed appropriate. If the I^2^ value is below 50%, a fixed-effect model will be used in the analysis. If the I^2^ is equal to or greater than 50%, heterogeneity will be considered high, and a random-effects model will be applied ^[20,21]^.

### 2.10 Subgroup or Subset Analysis

The choice of statistical model will be based on the assessment of heterogeneity: in the absence of substantial heterogeneity, a fixed-effect model will be used; if significant heterogeneity is observed (I^2^ ≥ 50%), a random-effects model will be employed to pool the results.

Subgroup analyses will be conducted when appropriate, considering factors such as different stingless bee species, animal models used, wound etiologies, and other relevant variables.

If eligible studies do not provide sufficient data for inclusion in the meta-analysis, their findings will be synthesized narratively and presented in tables summarizing the main characteristics and results.

### 2.12 Sensitivity Analysis

Sensitivity analyses will be conducted to assess the robustness of the conclusions by exploring the impact of methodological quality and sample size of the included studies. These analyses will only be performed if there are a sufficient number of studies to support a meta-analysis.

### 2.13 Publication Bias

To minimize publication bias, a comprehensive literature search will be conducted. To assess the presence of publication bias, Egger’s test will be applied. If necessary, the Duval and Tweedie’s trim-and-fill method will be used to adjust for potential effects of this bias on the results.

Both the forest plot and funnel plot will be analyzed to visualize data dispersion and assess publication bias, supporting the interpretation of the findings.

### 2.19 Ethics and Dissemination

As this study does not involve human subjects and will use only previously published data, ethical approval is not required.

The results of this review will be published in a peer-reviewed scientific journal relevant to the field of wound treatment. The authors are committed to disseminating the findings to the scientific community through national and international conferences, as well as through social media, digital media, and the websites of the university and research group, using accessible language aimed at both the general public and healthcare professionals.

## 3. Discussion

Stingless bee honey has been investigated as a therapeutic alternative for wound treatment, demonstrating important antimicrobial, anti-inflammatory, and wound-healing properties. However, a comprehensive and systematic evaluation of its clinical efficacy and safety is still needed to support its use across diverse healthcare settings [1].

Several factors may influence clinical outcomes related to the use of stingless bee honey, including its specific composition, the type of wound being treated, the method of application, and the clinical condition of the patient. Studies suggest that stingless bee honey has the potential to accelerate wound healing, reduce local infections, and improve overall wound quality ^[1,11]^.

More robust evidence on stingless bee honey may support the development of new clinical guidelines for wound management, particularly in vulnerable populations and low-resource settings. Future research should also take into account variables such as the geographic origin of the honey, differences in chemical composition among stingless bee species, and comparisons with conventional treatments, such as advanced wound dressings and topical medications.

In this context, further research into stingless bees and their honey—focusing on safety, standardization, methods of application, and evidence of efficacy—is essential for advancing scientific knowledge and developing new wound care therapies. Such progress could contribute to improved patient quality of life and the evolution of medical practice.

### 3.1 Strengths and limitations of this study

A comprehensive literature search will be conducted across 10 databases, following a rigorous protocol for data extraction and analysis. In addition, the included studies will be assessed using validated and widely accepted quality appraisal tools recommended in the scientific literature.

Among the potential limitations of this review is the likelihood of high heterogeneity across studies. This may be influenced by differences in the species of bees producing the honeys analyzed, the types of wounds treated, variations in animal models, assessment scales, and treatment protocols.

To mitigate the effects of high heterogeneity, heterogeneity will be evaluated and quantified using the I^2^ statistic (Inconsistency Index), and adjustments will be made to account for high heterogeneity when necessary.

Subgroup analyses will also contribute to reducing heterogeneity by enabling the examination of more homogeneous groups. Studies will be divided into smaller, more uniform subgroups based on relevant variables, such as: type of animal model used (e.g., rats, mice, rabbits), bee species involved in the study (*Melipona scutellaris, Melipona quadrifasciata*, etc.), type of wound treated (e.g., burns, ulcers, incisions), and the methodology employed to assess outcomes (e.g., healing, pain, infection).

## Data Availability

All relevant data from this study will be made available upon study completion.

## Acknowledgments

We would like to thank the educational institutions and the academic advisor involved in the development of this project.

